# Comparison of visual function analysis of people with low vision using three different models of augmented reality devices

**DOI:** 10.1101/2024.09.11.24313484

**Authors:** Sarika Gopalakrishnan, Arathy Kartha, Ronald Schuchard, Donald Fletcher

**Affiliations:** Envision Research Institute, USA; SUNY, Department of Biological and Vision Sciences, SUNY College of Optometry, New York, USA; Biotechnical R&D Consultant, USA; Envision Vision Rehabilitation Center, Wichita, Kansas; Smith-Kettlewell Eye Research Institute, San Francisco, California, USA

**Keywords:** Augmented reality, low vision, visual function, functional vision score, head-mounted display, contrast

## Abstract

We compared visual function in individuals with low vision (>20/60) using three different models of augmented reality (AR) devices: Ziru, IrisVision, and NuEyes-Pro3. The distance visual acuity (VA) was measured in high luminance high contrast (HLHC), high luminance low contrast (HLLC), low luminance high contrast (LLHC), and low luminance low contrast (LLLC) settings. The other tests were near VA, distance and near contrast sensitivity (CS), color vision, depth perception and indoor navigation. The change in visual function without and with AR devices was analyzed. Out of 27 participants, 17 were female. The mean age was 66.7±18.2 years. The median baseline VA was 0.66 (0.49) logMAR in HLHC, 0.87 (0.54) logMAR in HLLC, 0.84 (0.67) logMAR in LLHC and 1.04 (0.34) logMAR in LLLC. The median baseline near VA was 0.55(0.4) logMAR, distance and near CS was 1.10(0.26) logCS, and 1.20(0.30) logCS respectively. Distance and near vision showed significant differences with both Ziru and IrisVision (p<0.01), but not with NuEyes. There was a significant change in CS using Ziru and IrisVision for both distance and near (p<0.05) but both reduced significantly with NuEyes (p<0.01). The baseline functional vision score (FVS) was 45.76 (44.47) which improved to 79.04 (33.98) with Ziru and 76.14 (33.76) with IrisVision significantly, whereas it significantly reduced to 35.00 (33.97) with NuEyes (p<0.01). During the objective identification task on the indoor mobility course using AR devices, head-level objects were missed more compared to waist or floor-level objects across all three models. Majority of the visual functions improved with Ziru and IrisVision, with limited improvement in certain lighting condition of distance visual acuity with NuEyes.

## Introduction

Globally, an estimated 252.6 (95% CI, 111.4-424.5) million people live with low vision, i.e. a best-corrected visual acuity of 20/60 or worse in the better-seeing eye^[1]^. It is estimated that 7.08 million Americans have vision impairment, of whom 1.08 million people are living with blindness^[2]^. Ocular pathologies like diabetic retinopathy, glaucoma, age-related macular degeneration and other conditions can cause low vision^[5]^. Depending on the pathology, people with low vision would experience central field loss or peripheral field loss and /or generalized vision loss^[7]^.

People with low vision can have one or more decreased visual functions such as visual acuity, contrast sensitivity, color perception, depth perception and/or loss of visual field^[8]^. They may also have decreased functional vision such as difficulty in viewing distant objects, reading and writing tasks, independent mobility, photosensitivity, and performing household activities^9^. When these individuals find difficulty in visual tasks, sometimes they need to depend on caregivers for daily activities or they will stop doing certain tasks. Consequently, low vision can have a significant impact on their participation in activities of interest, independence, social interactions, quality of life, physical, emotional and mental health^[10–12]^. People fear losing vision more than memory, hearing, or speech, and consider visual loss among the top four worst things that could happen to them^[5]^. Rein et al estimated an economic burden of vision loss of $134.2 billion, those with vision loss incurred $16838 per year in incremental burden^[3]^. More than half of the visually impaired population do not even enter the workforce.

There is enough evidence that even though people with low vision have decreased form vision, they have some residual vision to perform certain range of visual activities^[6]^. Low vision rehabilitation services which include prescribing conventional optical devices and electronic devices are beneficial in improving the functional ability of people with low vision^[7]^. Yet, people face challenges when they need to use more than one low vision device for performing a variety of visual tasks at different distances for their daily living activities. To address these challenges, a range of assistive devices have been developed, including augmented reality (AR) head mounted displays which can be used for various visual tasks^[8,13]^.

Augmented reality refers to graphic overlays on, or graphic objects inserted in, live images of the real environment. Augmented reality head-mounted displays (AR-HMDs) enable users to see real images of the outside world and visualize virtual information generated by a computer at any time and from any location, making them useful for various applications. The manufacture of AR-HMDs combines the fields of optical engineering, optical materials, optical coating, precision manufacturing, electronic science, computer science, physiology, ergonomics, etc^[14,15]^. The basic principle of the AR device is to enhance the image cast by the world onto the retina with an enhanced view^[16]^. The cameras facing outward capture live video of the real world in front of the user, this video is processed to increase visibility via magnification and/or contrast enhancement and then shown in real-time to the user through a pair of micro-displays positioned in front of the eyes. This is called ‘video see-through display’ because even though the system is using a mobile phone, the users’ eyes are covered by opaque screens^[16,17]^.

Augmented reality devices are designed for different environments, both indoors and outdoors, and strive to be lightweight and comfortable to facilitate continuous and long-term wear. These devices are designed for use throughout the day while the user is engaged in other activities^[4,19]^. There are many AR devices developed to improve the visual function of people with low vision which include LVES, e-sight, Iris Vision, Hololens, Oxsight, NuEyes, Vision Buddy, etc.^[18]^. In previous studies, there is some information on one or two visual functions assessed with a single virtual reality (VR) or augmented reality (AR) device. However, there are no studies that compared different models of AR devices. Few studies reviewed head-mounted displays (HMDs) for visual impairment, highlighting their advantages over conventional desk-mounted or handheld low vision devices^[16,21-25]^. Crossland et al. suggested that 47% of the study participants were ready to use a device like Sightplus^25^, while Yeo et al reported 94% of the participants were ready to use similar device called Relumino^22^. Htike et al. reported that Hololens was useful for navigational task, depth perception and object recognition tasks^25^ and Dylan et al. showed that augmentation helps in obstacle identification^26^. However, no study has compared the depth perception, object identification, or navigation tasks among different models of AR devices, which would provide varying experiences with different device models.

While there are different generations of AR devices and a variety of models, it is important to examine how the visual abilities of people with low vision vary while using different models of AR devices. While these systems seem promising, there is less evidence on comparing different models of these devices in improving the visual functions of people with low vision^[18]^. Therefore, to understand the visual abilities of people with low vision, we aim to explore what and how they could use different models of AR devices in both stationary and mobile scenarios. We address the following research questions in this paper: 1. What is the change in visual function while using three models of AR devices? 2. Which model of AR device helps better for which type of functional vision? 3. How do AR devices help in identifying objects while moving in an indoor environment?

## Methods

This was a prospective study design, and all study participants were recruited between 02/01/2023 and 31/05/2023 from the participant database of the Envision Research Institute and Envision Vision Rehabilitation Center.

### Participants

Participants with low vision who had best corrected visual acuity of worse than 20/60, central field loss (CFL), or peripheral field loss (PFL) were recruited for the study. Those with cognitive impairment, physical disabilities, and/or multiple disabilities were excluded. The informed consent form explained the study protocol, time commitment, risks, and benefits to the participants. All the participants provided written consent for participation before the study. The study adhered to the tenets of the Declaration of Helsinki, and it was approved by the institutional review board and ethics committee of Envision Research Institute and Wichita State University, USA.

### Procedure

The visual functions were evaluated without any AR device with the participant’s best refractive correction. The concept of augmented reality technology was explained to the participants and a detailed demonstration of all three models of AR devices was given before starting the experiment. They were trained to adjust their desired magnification level, viewing/ contrast mode, and other functions of the devices. All the visual tasks were measured with all three models of AR devices in a randomized order.

### AR devices

We used three models of augmented reality devices to analyze the visual functions and functional vision of people with low vision in this study. The devices were, a) Ziru (Snapdragon model, Dodrutu, Oxford, UK) a smartphone Samsung Galaxy S9 fitted on a 3D printed hardware case that is fixed on a golf cap, b) Iris Vision (Iris Vision live model, IrisVision Global, Inc. CA, USA) provides a display on a smartphone Samsung S10 fitted on a Samsung gear VR headset and c) NuEyes (NuEyes Pro3, NuEyes Technologies Inc., CA, USA) is a spectacle model device with a display which is connected to a smartphone Samsung S20 model. The features of all three devices are listed in Table 1.

**Table 1:**
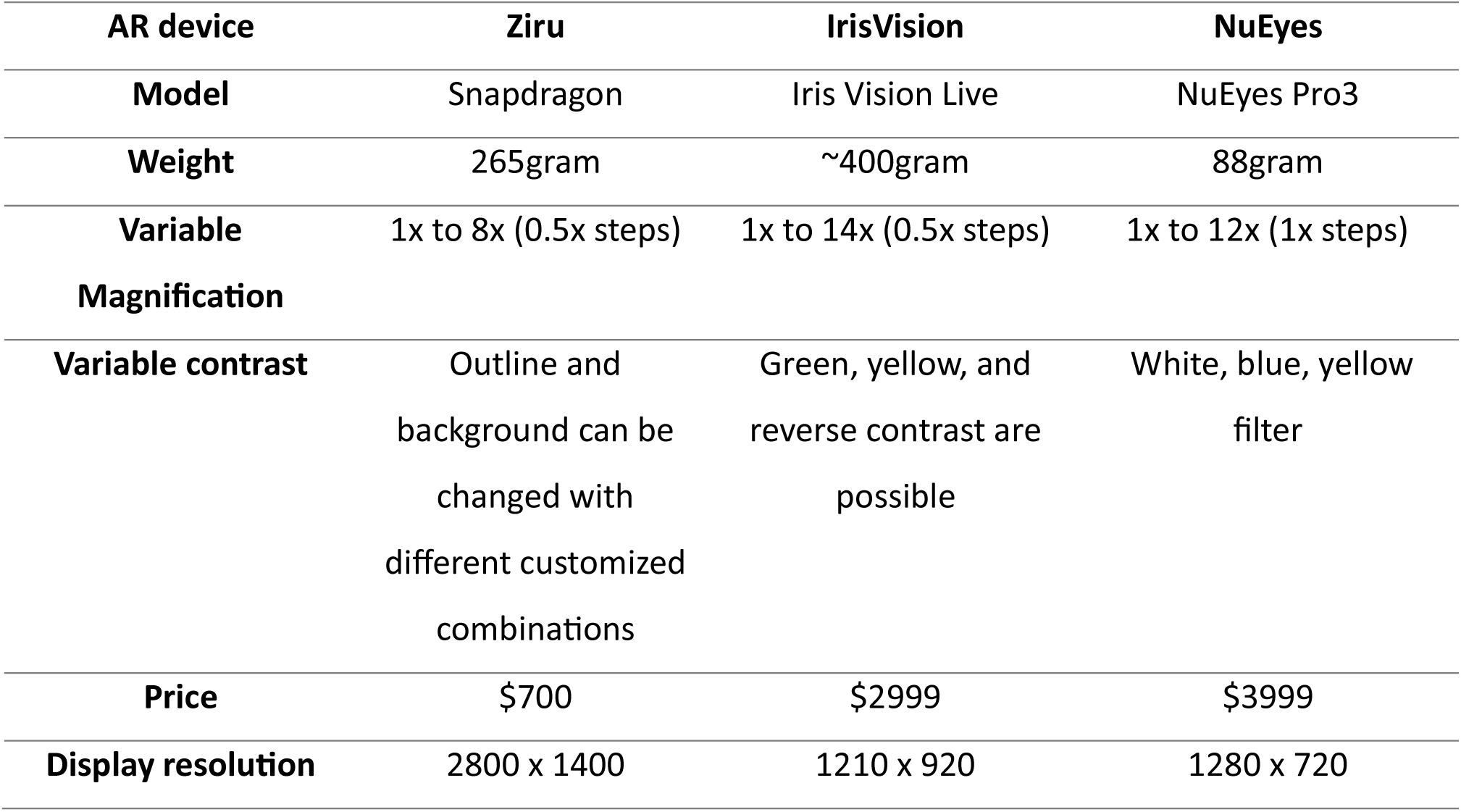
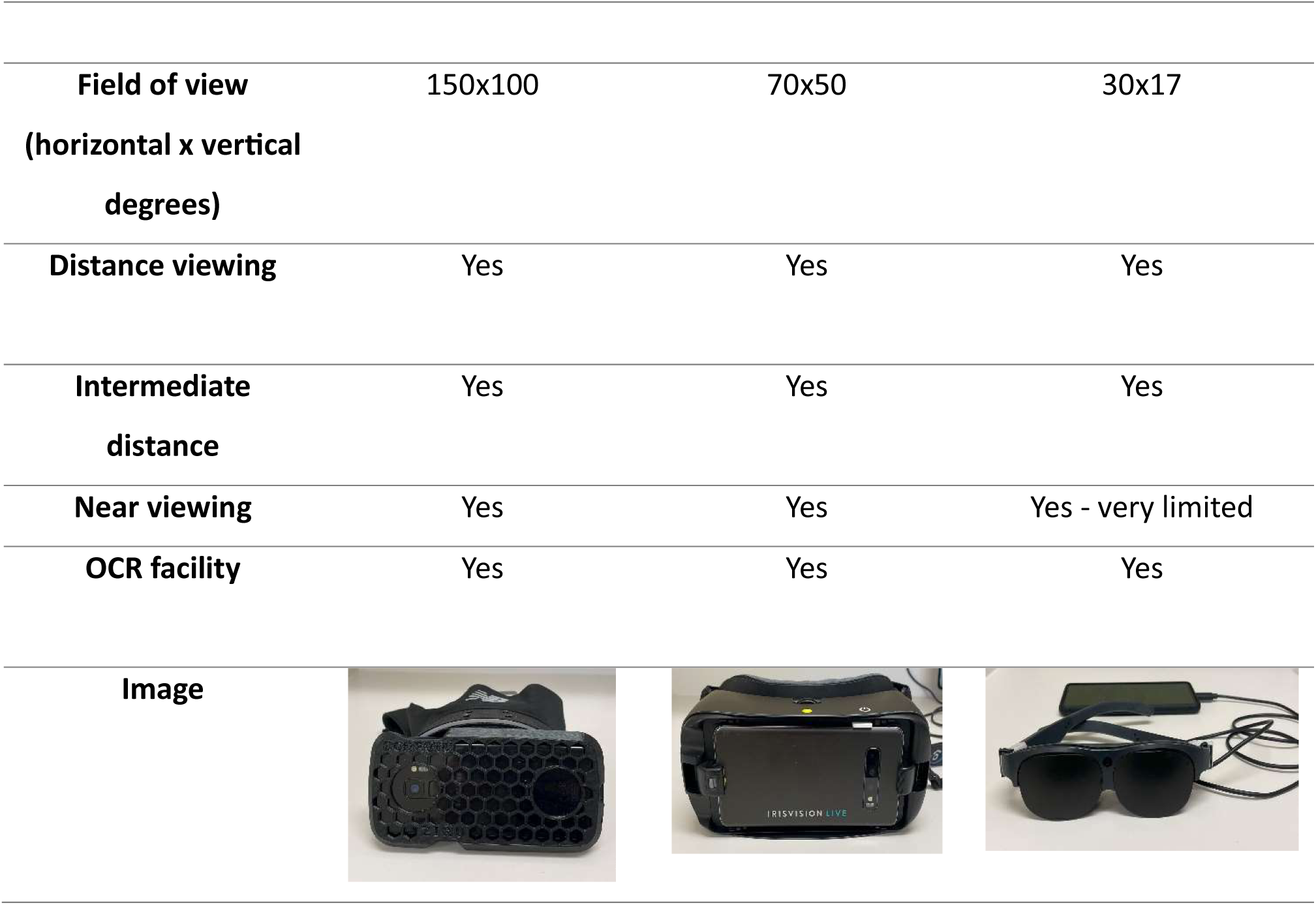
Technical specifications of the augmented reality devices.

| **AR device** | **Ziru** | **IrisVision** | **NuEyes** |
| --- | --- | --- | --- |
| **Model** | Snapdragon | Iris Vision Live | NuEyes Pro3 |
| **Weight** | 265gram | ~400gram | 88gram |
| **Variable Magnification** | 1x to 8x (0.5x steps) | 1x to 14x (0.5x steps) | 1x to 12x (1x steps) |
| **Variable contrast** | Outline and background can be changed with different customized combinations | Green, yellow, and reverse contrast are possible | White, blue, yellow filter |
| **Price** | \$700 | \$2999 | \$3999 |
| **Display resolution** | 2800 x 1400 | 1210 x 920 | 1280 x 720 |
| **Field of view**<br>(horizontal x vertical<br>degrees) | 150x100 | 70x50 | 30x17 |
| **Distance viewing** | Yes | Yes | Yes |
| **Intermediate**<br>**distance** | Yes | Yes | Yes |
| **Near viewing** | Yes | Yes | Yes - very limited |
| **OCR facility** | Yes | Yes | Yes |
| **Image** |  |  |  |

Ziru device provides up to 8 times magnification with five different viewing modes such as ‘CLEAR’-color lens designed to reproduce true colors and object proportions and sizes, ‘BOOST’-composite lens combining CLEAR lens output with OUTLINE lens, ‘OUTLINE’-lens that filters out everything but object outlines, ‘COGNI’-composite lens combining CLEAR lens with a number of other lenses designed to enhance visual cognition, and ‘GREY’-gray scale lens with adaptive brightness and contrast. The graphical representation of the viewing modes can be seen in Supplementary figure (S1 Fig). It is a head-mounted display that has autofocus capacity from near to far distance and image stabilization processing which will help match the head movement of the user along with the device movement. It has a low-latency video pipeline with a 4K camera system and a 2K display system. The user can wear his or her spectacle prescription while wearing this device. It accepts voice commands from users to change the viewing modes, brightness, magnification, and other most frequently used functions, in addition to that, the magnification level can be adjusted using a physical button on top of the device or through voice commands. It also has optical character recognition (OCR) function which helps user to read out any printed material, sometimes it can read handwritten material. Ziru can be calibrated for each user, and it has the feature of adjustable interpupillary distance, therefore it can be used from a young age to elderly people. It does not need an internet or Wi-Fi connection for it to work, which makes it more functional in a wide variety of environments.

IrisVision provides up to 14 times magnification with 10 different viewing modes such as, ‘SCENE’-regular mode, ‘SCENE WITH BUBBLE’-regular mode with additional magnified area in the shape of a bubble, ‘RP’ - scene mode with a reduced field of view, ‘TELEVISION’ - designed specifically for watching television, ‘BIOPTIC’ - scene mode with an additional magnified view in a rectangle-shaped box, ‘READING’ - Magnified version designed for reading tasks, ‘READING INVERTED’-reading mode with reverse contrast as white text on black background, ‘READING LINE’ - reading mode with a line to fixate, ‘READING GREEN’ - green text on black background AND ‘READING YELLOW’-yellow text on black background. The representation can be viewed in Supplementary Fig.B. The head-mounted display uses a touch panel to specify operations to be performed. It has features of contrast enhancement, image remapping strategies, motion compensation strategies, augmented reality strategies, and OCR functions. It does need a network connection to update the software regularly.

NuEyes Pro3 provides up to 12 times magnification with five color filters such as ‘Black’ - Black on white, ‘White’ - white on black, ‘Yellow’ - yellow on black, ‘Orange’ - orange on black, and ‘Blue’ - blue on yellow apart from the regular viewing mode as showed in Supplementary Fig.C. It presents three resolution levels high, medium, and low. The spectacle display has an extended wire that connects to a smartphone on which the operations are performed. The user has an option of fitting an additional clip-on lens in front of the camera on the spectacle to focus objects at a far distance.

The luminance on the visual acuity chart was measured using Konica Minolta Spot Photometer F (Digital Light Exposure Spot Meter, JAPAN) on a uniform background in a controlled environment without and with all three AR devices. Ten recordings were taken in cd/m^2^ to measure the luminance on the display screen of each of the AR devices. The background room illumination was measured in lux which was noted to be 273.4 lux for high luminance conditions and 0.01 lux for low luminance conditions.

### Visual functions

The distance visual acuity was measured with internally illuminated ETDRS high and low contrast 4m logMAR charts. The visual acuity was evaluated in four conditions, i) High luminance high contrast (HLHC), ii) High luminance low contrast (HLLC), iii) Low luminance high contrast (LLHC), and iv) Low luminance low contrast (LLLC). The high and low luminance levels used on the illuminated cabinet were 160 cd/m^2^ and 3 cd/m^2^ respectively. The contrast levels used were 5%, 10% and 25% for low contrast conditions and 100% for high contrast conditions. Initially, the participants were tested with 5% contrast logMAR chart for low contrast condition, if they were not able to respond to 5%, then 10% contrast chart was tried, and if they had difficulty responding to that, then 25% contrast chart was used. The participants were encouraged to adjust the level of magnification until they were able to recognize the smallest possible optotypes on the logMAR chart. The near visual acuity was tested with the SK Read chart at their habitual reading distance. The distance and near contrast sensitivity were evaluated using distance and near Pelli Robson contrast sensitivity charts respectively. The central field score was assessed using the Berkeley central field test and the peripheral field score was assessed using tangent screen. The color vision was evaluated using the PV-16 test, stereopsis was measured using the Frisby test.

### Functional vision score

#### Functional acuity score

Every logMAR value of visual acuity has an equivalent visual acuity rate (VAR) value which is known as visual acuity score (VAS). It can be calculated using the formula, VAS = 100 - 50*logMAR. The VAS for the right eye (OD), left eye (OS), and both eyes (OU) were documented. The weighted average of three VAS was used to calculate the functional acuity score (FAS) according to the formula,

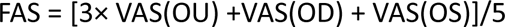

#### Functional field score

Out of 50 points, the number of points seen in the central field was marked as central field score (CFS), and in the peripheral field was documented as peripheral field score (PFS). The visual field score (VFS) was calculated as VFS = CFS + PFS for the right eye, left eye, and both eyes. The weighted average was calculated as functional field score (FFS) using the formula^[20]^,

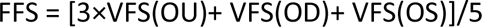

The functional vision score (FVS) was derived by multiplying the FAS and FFS using the formula,

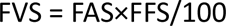

An FVS score of 100 or greater represents normal vision and a score of zero would represent total blindness.

### Depth perception task

Depth perception was evaluated with the help of a setup designed for this study. Five equally sized (L x W x H = 5 x 5 x 5 inch ∼12.7cm) cube-shaped boxes were suspended from the ceiling of the lab using clear fishing line at different distances from the participant’s viewing point (Fig 1). The boxes were labeled with random numbers in no specific order and were separated by 1 feet (∼0.3m) distance from each other box. The participant was positioned at 12 feet (∼3.7m) from the closest box. The top view of the scene was obscured using black charts so that the fishing lines connecting the boxes to the ceiling were not visible to the participants, to avoid any additional depth cues. Using a 2-alternative forced choice, each participant was asked to look at two boxes in a pairwise combination, and the participant was asked to respond which box was seen closer when compared to the other box. Likewise, ten sets of combinations were presented to the participants based on which ten responses were received from each participant. Participants performed the task without and with all three AR devices.

**Figure 1.**
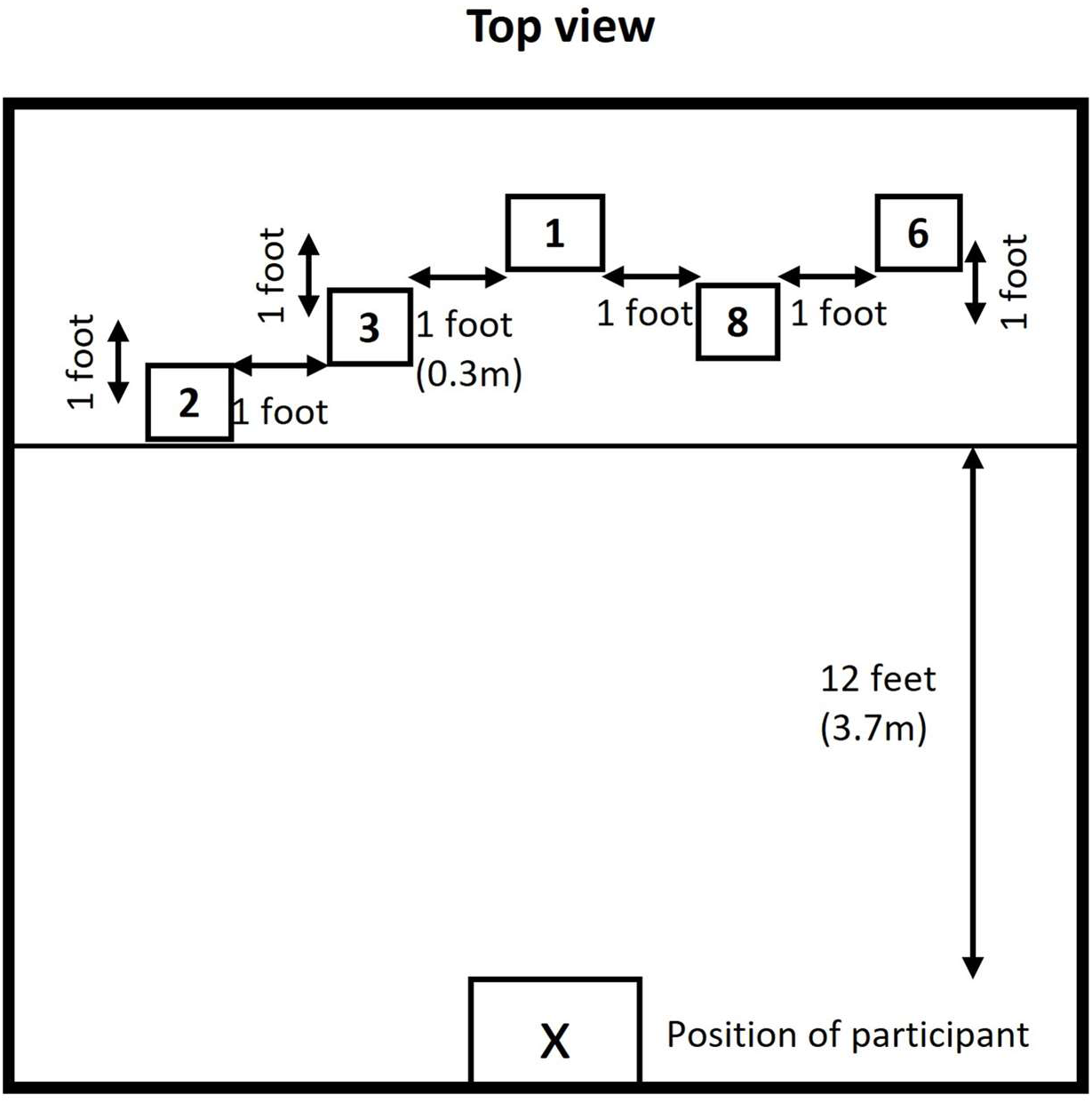
Depth perception test.

Fig 1 Description: X - Position of the participant, five equal-sized cube-shaped boxes were hanging from the room ceiling at different distances from the participant.

### Object identification task

The object identification task was done on a mobility course designed in a quiet room (20×20 feet ∼6m). The center of the room had four black curtains that were used to create a cross-shaped barrier, therefore the participants had four quadrants of space for them to explore. This barrier facilitated obscuring other quadrants of the room so that the participants could not see all the objects at the same time. The participants were instructed to walk at their usual walking speed following the directions given in the room. The participants were supposed to identify the ten toys (soft or stuffed or plastic toys) positioned at floor level, waist level, and ceiling level while finding their way to the exit point. They had to identify a toy, point it out, and describe it shortly as what type of toy, what color, and what is the material of the toy in a few words. There were distractors like boxes, drums, and chairs in the room and pictures on the wall and curtain (all distractors were not toys) in all levels which added an adequate level of cognitive load to the task. Four different mobility courses were designed by changing the position of the toys at different levels and the walking direction (Fig 2). Initially, the task was performed without any AR device and then it was repeated with all three augmented reality devices in a randomized order. The time taken, accuracy, number of objects identified, number of objects missed at each level, and feedback from participants were documented. Participants were allowed to use their habitual cane for the task and they were accompanied by the experimenter at all times to ensure safety while walking.

**Figure 2.**
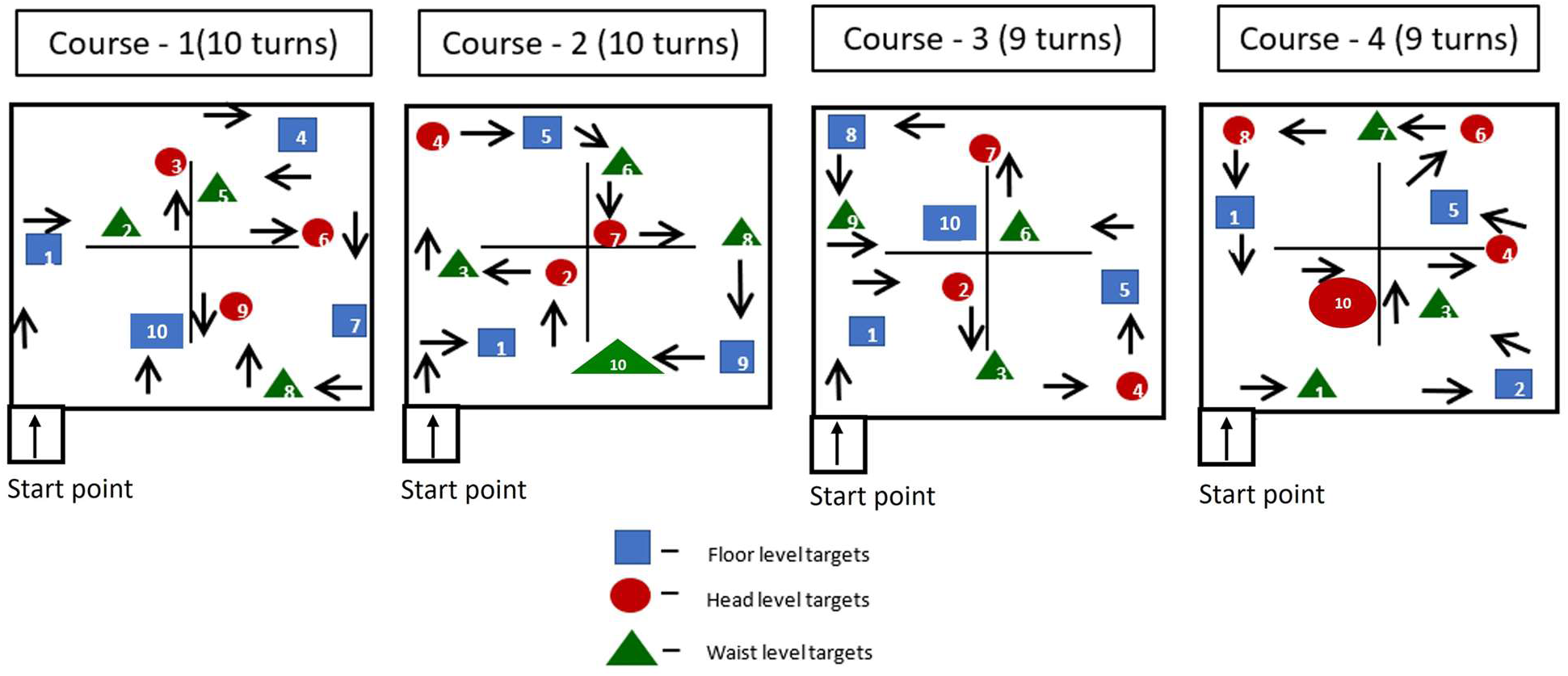
Representation of mobility course for object identification task.

Fig 2 Description: The participant entered the mobility course at the entrance point, followed the direction signs, identified the objects at head level, waist level and floor level and after completing the task came out of the mobility course through the same entrance point.

### Statistical Analysis

Statistical analyses were conducted using SPSS software version 20. Descriptive measures, including participants’ demographic and clinical characteristics, were summarized as means and standard deviations, and medians and interquartile ranges, and by counts and percentages as appropriate. The repeated measures of the AR devices were analyzed using the Friedman test and the Wilcoxon sign rank test was performed to measure the differences between groups. The alpha level was set at 0.05.

## Results

Out of 27 patients, 17 (63%) were female. The mean age of the participants was 66.7 (SD, 18.2) ranging from 25 to 92 years of age, with the majority of them (40.7%, n=11) ranging from 61 to 80 years of age. Thirteen people were noted to have ocular pathology causing central field loss, 9 had peripheral field loss, 2 of them had central and peripheral field loss and 3 had generalized vision loss. Most of them, 55.6% (n=15), were retired, 25.9% (n=7) were artists and the rest 18.5% (n=5) were involved in private jobs. Nearly half of them, 48.1% (n=13), had physical morbidities, 22.2% (n=6) had hearing impairment and 48.1% (n=13) were using mobility assistance. More than half of them 14 (51.9%) had previous experience using low vision devices. Most of them, (44.4%, n=12), had a duration of visual impairment of less than 10 years, followed by 10 to 20 years (22.2%, n=6) as shown in Table 2. Mean results with standard deviations in parenthesis are reported for the measurements. Statistically significant differences were determined at p<0.05.

**Table 2:**
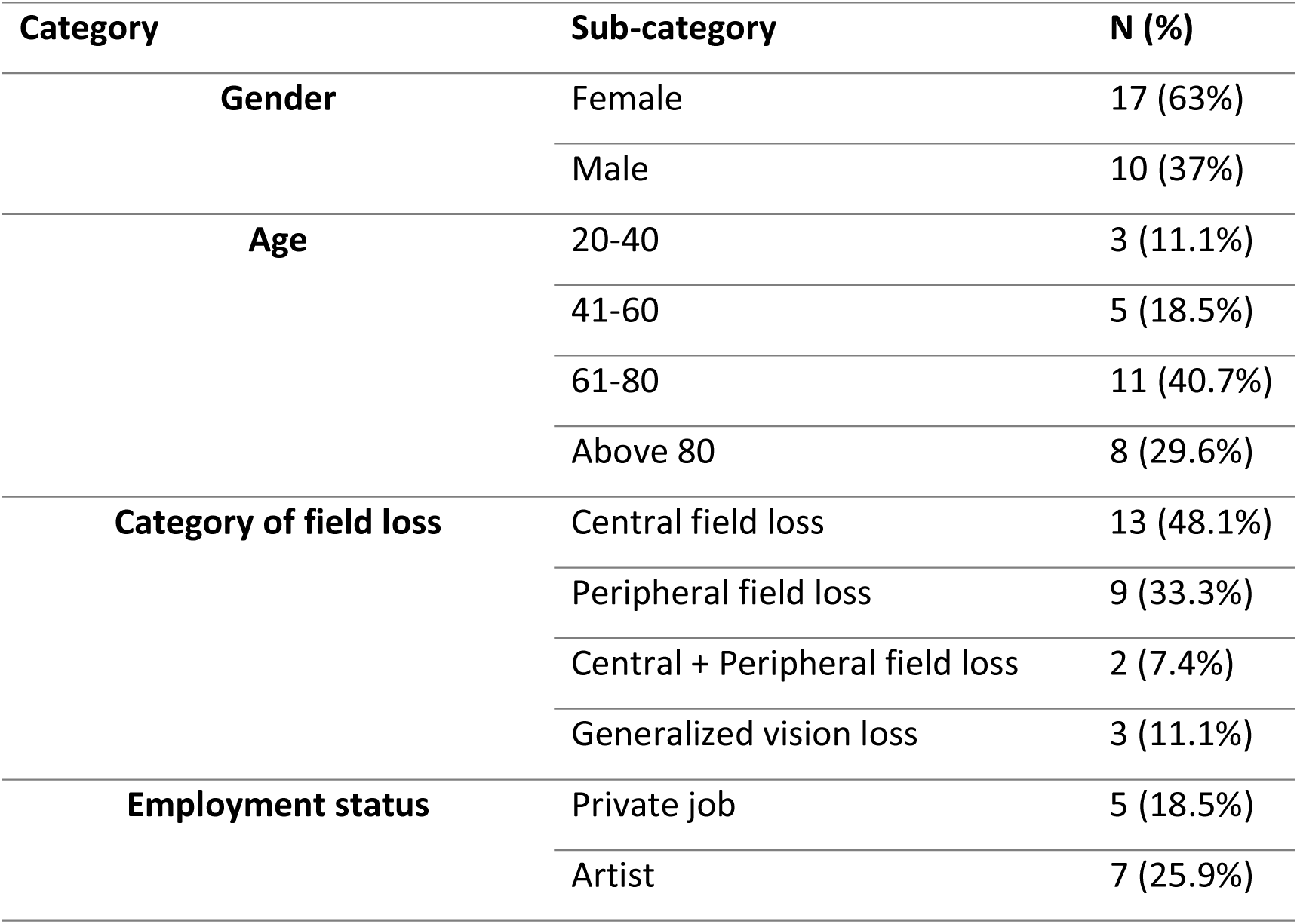
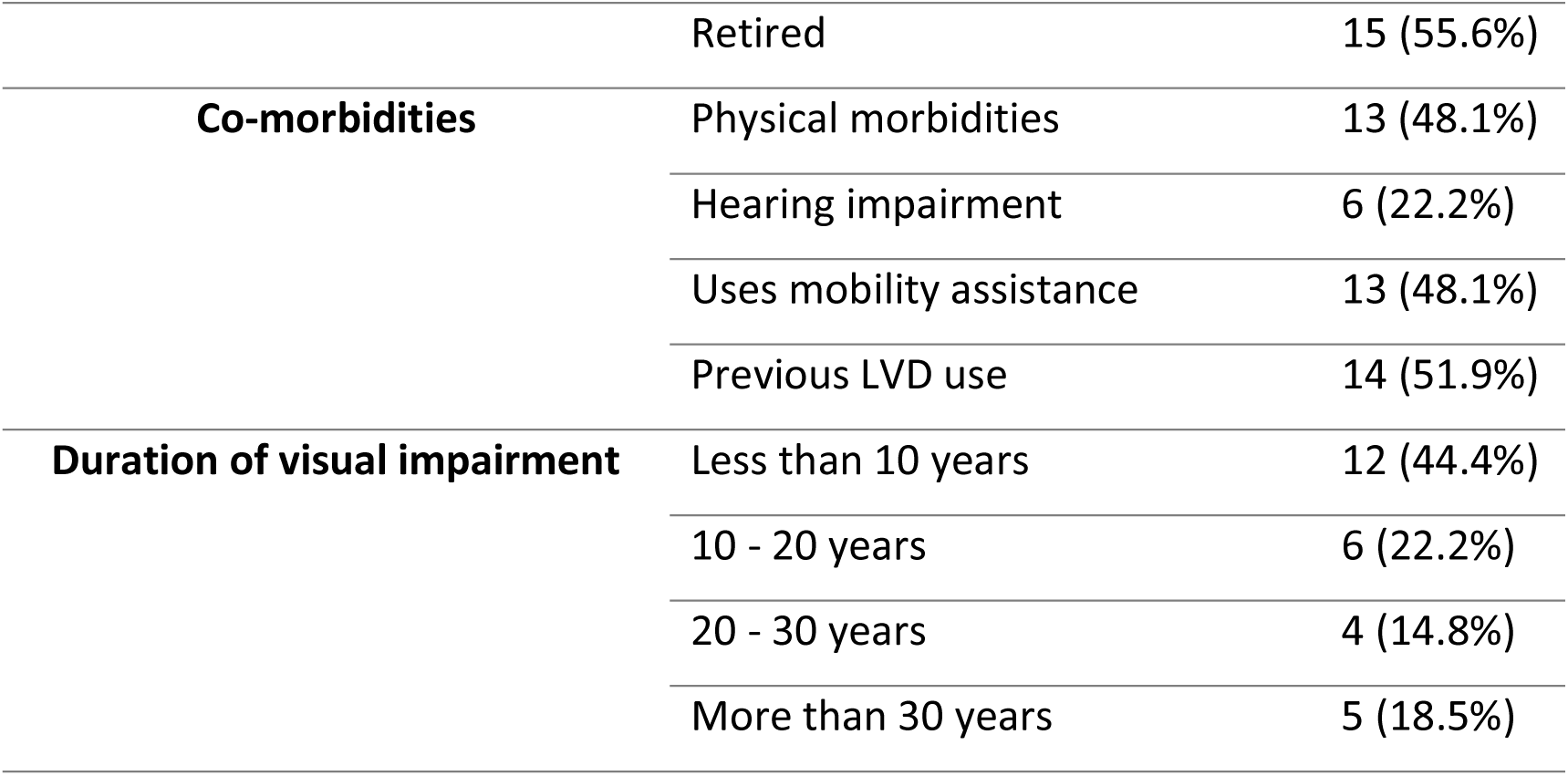
Demographic characteristics of study participants.

There was a statistically significant difference in the number of lines read without and while using three models of augmented reality devices on all the lighting conditions, HLHC χ^2^_(3)_=60.26, p<0.0001, HLLC χ^2^_(3)_=16.42, p=0.001, LLHC χ^2^_(3)_=53.25, p<0.0001, LLLC χ^2^_(3)_=10.57, p=0.014. Post hoc analysis with Wilcoxon signed-rank tests was conducted with Bonferroni correction applied, resulting in a significance level set at p<0.05. The median (IQR) distance visual acuity in HLHC was 0.66 (0.49) logMAR in the HLHC condition and they could read 0.20 (0.44) logMAR with Ziru with a maximum of 8x magnification (Z = −4.541, p<0.01) and 0.16 (0.25) logMAR with Iris vision with a maximum magnification of 14x (Z = −4.469, p<0.01), but with NuEyes, the median visual acuity of 0.74 (0.47) logMAR was not significantly different when compared to the presenting visual acuity (Z=-1.562, p=0.12). The median distance visual acuity in HLLC was 0.87 (0.54) logMAR and they could read 0.61 (0.32) with Ziru (Z=-3.125, p=0.002) and 0.64 (0.40) with Iris vision (Z=-2.627, p=0.009) but with NuEyes, it was 0.82 (0.34) logMAR which was not significantly different (Z=-1.014, p=0.311). The median distance visual acuity in LLHC condition was 0.84 (0.67) logMAR, they were able to read 0.24 (0.19) with Ziru (Z=-4.458, p<0.01), 0.24 (0.29) with Iris Vision (Z=-4.542, p<0.01), and 0.60 (0.44) with NuEyes (Z=-3.036, p=0.002), the number of lines read were significantly different with all the three AR devices when compared to the presenting visual acuity. The median visual acuity in the LLLC condition was 1.04 (0.34) logMAR, and they were able to read 0.70 (0.32) with Ziru (Z=-3.365, p=0.001), 0.68 (0.37) with Iris vision (Z=-2.772, p=0.006) and 0.94 (0.26) with NuEyes (Z=-2.499, p=0.012), which was statistically significant with all three devices.

The mean luminance without AR devices was measured as 73.8 (9.1) cd/m^2^, it was 76.4 (11.6) cd/m^2^ with Ziru which was not significantly different (p=0.47) when compared to luminance without any AR device, but it was 13.3 (0.6) cd/m^2^ with IrisVision and 6.2 (3.7) cd/m^2^ with NuEyes which were significantly different (p<0.05). The luminance was significantly different among all the three devices (p<0.01). On HLLC condition, more than half of the participants 60% were able to read the chart at 25% contrast, 24% of them read at 10% contrast and 12% of them read at 5% contrast without any AR device. Interestingly, a maximum number of participants were able to read 5% low contrast level chart with Ziru (28%), followed by IrisVision (16%), none of the participants were able to read the 5% contrast level chart with NuEyes. With IrisVision, most of them 64% were able to respond to 10% contrast chart and with NuEyes, 40% of them could respond to 25% contrast level. In case of LLLC condition, 60% of them read the chart at 25% contrast, 16% read at 10% contrast and 12% read at 5% contrast level. Similar to the previous situation, the maximum number of participants read 5% low contrast level with Ziru (28%), followed by IrisVision (20%), none of them were able to read the 5% chart with NuEyes. Using IrisVision, 60% of them were able to read the chart at 10% level and with NuEyes, 32% were able to read at 25% contrast level.

There was a statistically significant difference in the near visual acuity without and while using three models of augmented reality devices, χ^2^_(3)_=51.17, p<0.0001. Post hoc analysis with Wilcoxon signed-rank tests was conducted resulting in a significance level set at p<0.05. The median (IQR) near visual acuity was 0.55 (0.40) logMAR, they could read 0.10 (0.30) logMAR with Ziru (Z=-4.376, p<0.0001) and 0.10 (0.30) logMAR with IrisVision (Z=-4.375, p<0.0001) which was significantly different with a p-value of <0.05, while it significantly reduced to 0.80 (0.20) logMAR with NuEyes (Z=-2.593, p=0.01). There was a statistically significant difference in the distance contrast sensitivity χ^2^_(3)_=38.36, p<0.0001 and near contrast sensitivity χ^2^_(3)_=45.42, p<0.0001 without and while using three models of augmented reality devices. The median (IQR) distance contrast sensitivity was 1.10 (0.26) logCS without the AR device, it was 1.20 (0.33) with Ziru (Z=-1.832, p=0.01), and 1.16 (0.43) with IrisVision (Z=-0.750, p=0.04) which was significantly different and it reduced significantly to 0.75 (0.45) with NuEyes (Z=-3.519, p<0.001). The median (IQR) near contrast sensitivity was 1.20 (0.30) logCS without using any AR device, it was 1.43 (0.41) with Ziru (Z=-2.589, p=0.01) and 1.35 (0.30) with Iris vision (Z=-2.267, p=0.02) which was significantly different (p<0.05) and it reduced significantly to 0.75 (0.64) with NuEyes (Z=-3.866, p<0.001) as shown in Table 3.

**Table 3:**
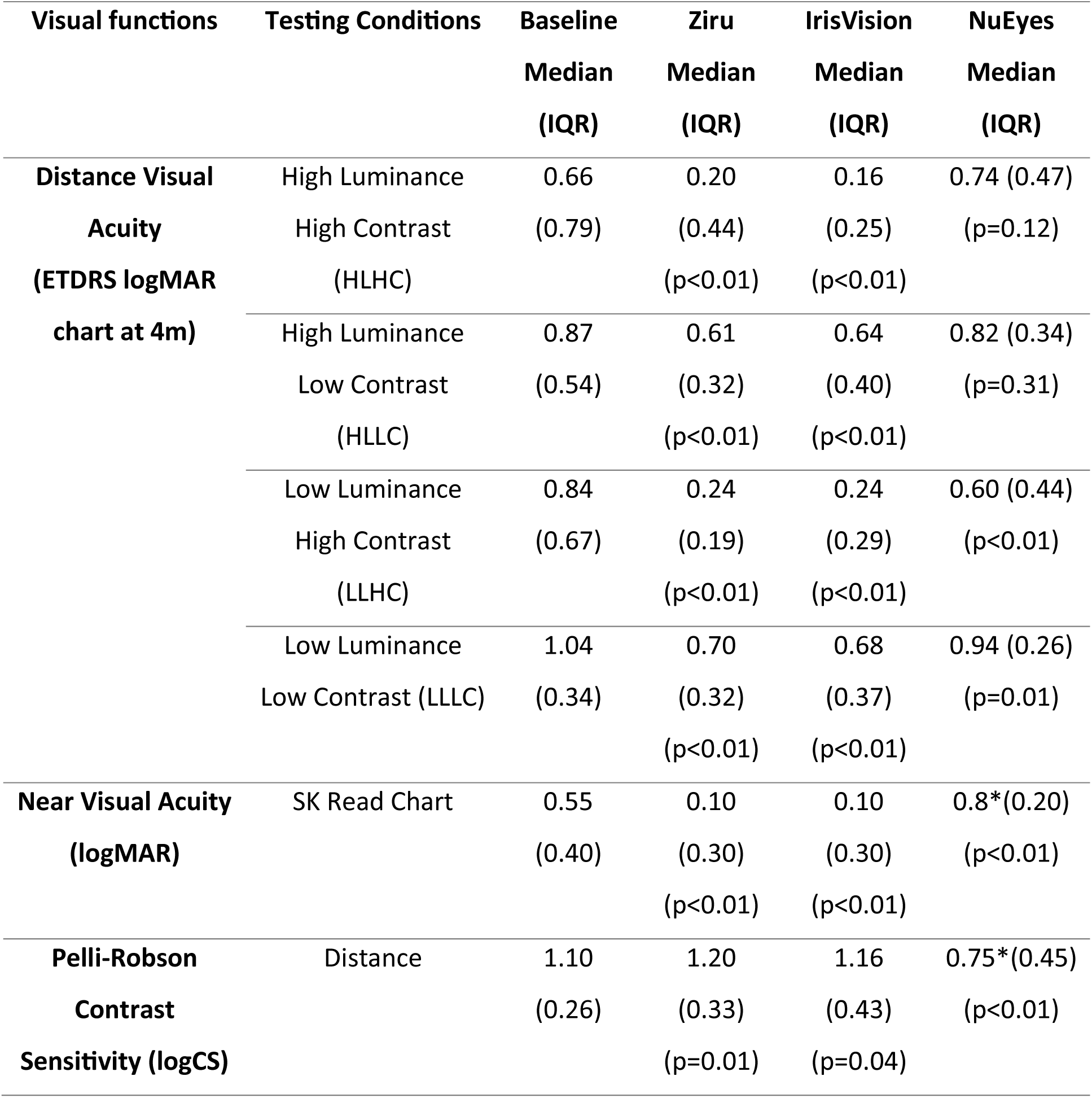
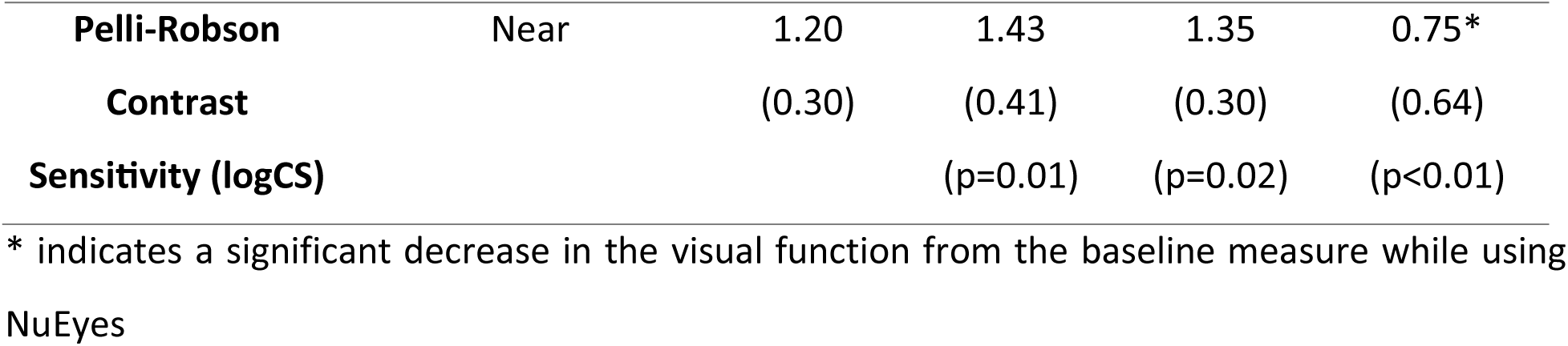
Visual functions of people with low vision while using three different models of augmented reality devices.

The mean correct response of the depth perception task was 46.2% without using any AR device, 50.4% with Ziru, 48.8% with Iris Vision, and 48.8% with NuEyes. In the PV-16 color vision test, 37% of the participants arranged the color caps without any errors, and this result was not altered by any of the AR devices. During the baseline assessment, 40.7% of the participants made major errors and 22.2% made minor errors. With Ziru, there were 25.9% major errors and 37% minor error, for IrisVision, the numbers were 44.4% major errors and 18.5% minor errors and with NuEyes, the findings were 29.6% major errors and 33.3% minor errors. No specific color vision deficiency with any viewing condition was noted for any participant. Three participants were able to respond to a 6mm Frisby plate and their stereo acuity was 215, 340, and 600 arc sec without any AR device, of which one participant responded to 3mm and 1.5mm with 170 and 55 arc sec respectively. There was no difference noted in stereo acuity with any of the AR devices when compared to without AR devices.

### Functional vision score

There was a statistically significant difference in the functional vision score without and while using three models of augmented reality devices, χ^2^_(3)_=54.37, p<0.0001. The median (IQR) functional acuity score (FAS) was 59.00 (37.40) which significantly improved to 90.00 (21.75) with Ziru, 92.00(12.50) with Iris Vision, and 63.00 (23.50) with NuEyes as shown in Table 4. The FAS improvement between Ziru and Iris Vision was not significantly different, however, the difference was significant when compared to NuEyes. The median functional field score was 86.00 (35.00) without the AR device, 82.00(28.00) with Ziru, 81.00 (31.50) with Iris vision but the functional field score was reduced significantly to 60.00 (37.00) with NuEyes, which could be due to the display size of the device which significantly reduced the peripheral field of view. However, the median composite functional vision score 45.76 (44.47) had significant improvement to 79.04 (33.98) with Ziru (Z=-4.469, p<0.0001) and 76.14 (33.76) with Iris Vision (Z=-4.228, p<0.0001), but the functional vision score reduced significantly with NuEyes to 35.00 (33.97) (Z=-2.811, p=0.005).

**Table 4:** Comparison of functional vision score of individuals with low vision while using three different augmented reality devices.

| Score | Baseline<br>Median<br>(IQR) | Ziru<br>Median (IQR) | IrisVision<br>Median (IQR) | NuEyes<br>Median (IQR) |
| --- | --- | --- | --- | --- |
| Functional acuity<br>score (FAS) | 59.00<br>(37.40) | 90.00 (21.75)<br>($p<0.01$) | 92.00 (12.50)<br>($p<0.01$) | 63.00 (23.50)<br>($p<0.01$) |
| Functional field<br>score (FFS) | 86.00<br>(35.00) | 82.00 (28.00)<br>($p=0.51$) | 81.00 (31.50)<br>($p=0.56$) | 60.00 (37.00)<br>($p<0.01$)* |
| Functional vision<br>score (FVS) | 45.76<br>(44.47) | 79.04 (33.98)<br>($p<0.01$) | 76.14(33.76)<br>($p<0.01$) | 35.00 (33.97)<br>($p<0.01$)* |

FAS: Functional acuity score, FFS; Functional field score, FVS: Functional vision score, \*p-value indicates a significant decrease in the visual function from the baseline measure

### Object identification task

All participants were able to complete the navigation task without and with all models of AR devices. Two participants reported mild symptoms of motion sickness while using IrisVision which was not reported while using Ziru or NuEyes. Out of 10 objects that were positioned at different heights, the participants were able to identify most of the objects using AR devices. The average number of objects identified was 9.43 (1.12) without an AR device which was not significantly different from any of the AR devices. The average time taken for completion of this task was not significantly different among all four conditions. The number of objects missed without using any AR devices and while using Ziru was not significantly different (p>0.05), but it was significantly different while using IrisVision and NuEyes (p<0.05) as shown in Table 5.

**Table 5:**
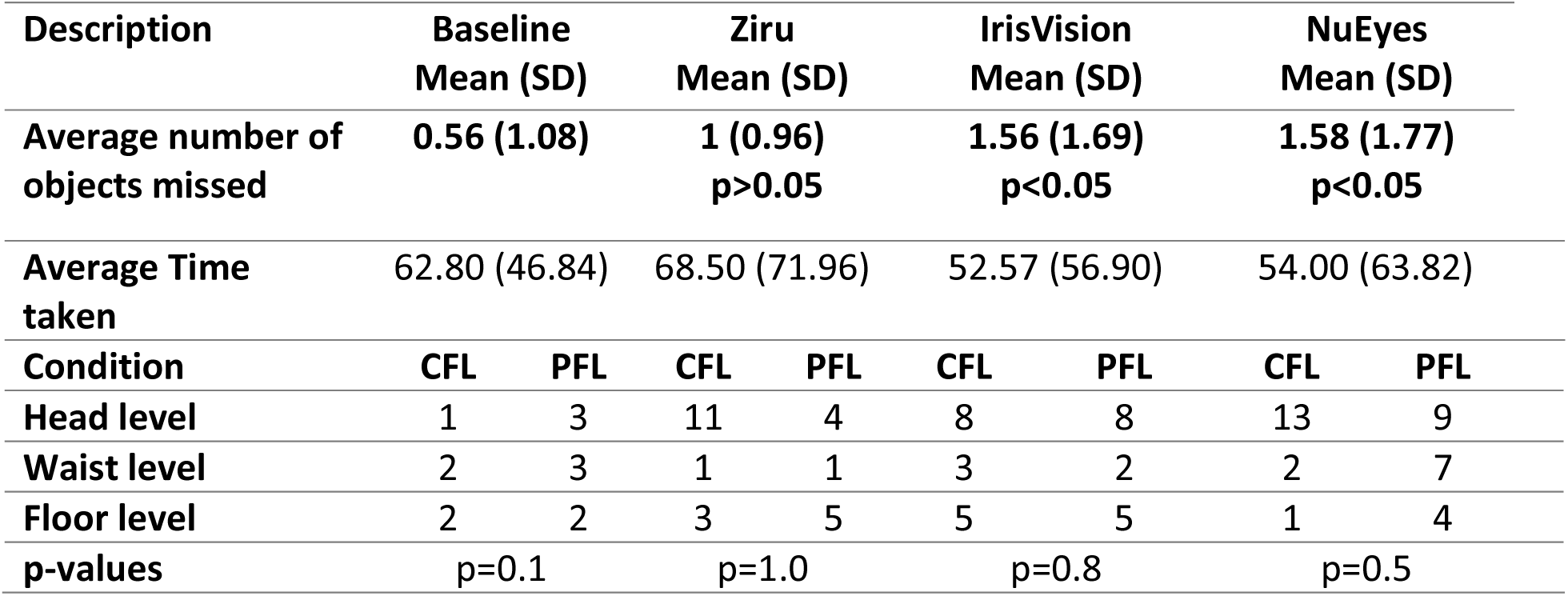
Object identification task using augmented reality devices.

| Description | Baseline<br>Mean (SD) |  | Ziru<br>Mean (SD) |  | IrisVision<br>Mean (SD) |  | NuEyes<br>Mean (SD) |  |
| --- | --- | --- | --- | --- | --- | --- | --- | --- |
| Average number of objects missed | 0.56 (1.08) |  | 1 (0.96)<br>p>0.05 |  | 1.56 (1.69)<br>p<0.05 |  | 1.58 (1.77)<br>p<0.05 |  |
| Average Time taken | 62.80 (46.84) |  | 68.50 (71.96) |  | 52.57 (56.90) |  | 54.00 (63.82) |  |
| Condition | CFL | PFL | CFL | PFL | CFL | PFL | CFL | PFL |
| Head level | 1 | 3 | 11 | 4 | 8 | 8 | 13 | 9 |
| Waist level | 2 | 3 | 1 | 1 | 3 | 2 | 2 | 7 |
| Floor level | 2 | 2 | 3 | 5 | 5 | 5 | 1 | 4 |
| p-values | p=0.1 |  | p=1.0 |  | p=0.8 |  | p=0.5 |  |

However, head-level objects (n=16,21,23 with Ziru, IrisVision and NuEyes) were missed more when compared to waist and floor levels while using any of the AR devices (Fig 3). This could be because of the field of view limitations in head-mounted displays. While wearing the AR headsets, the number of objects missed at head level was equal or higher among participants with CFL when compared to those with PFL, this could be because head-level objects are visualized using the central field of vision. Similarly, the number of objects missed at floor level was equal or higher among those with PFL when compared to those with CFL. However, the differences between the CFL and PFL did not reach statistical significance.

**Figure 3.**
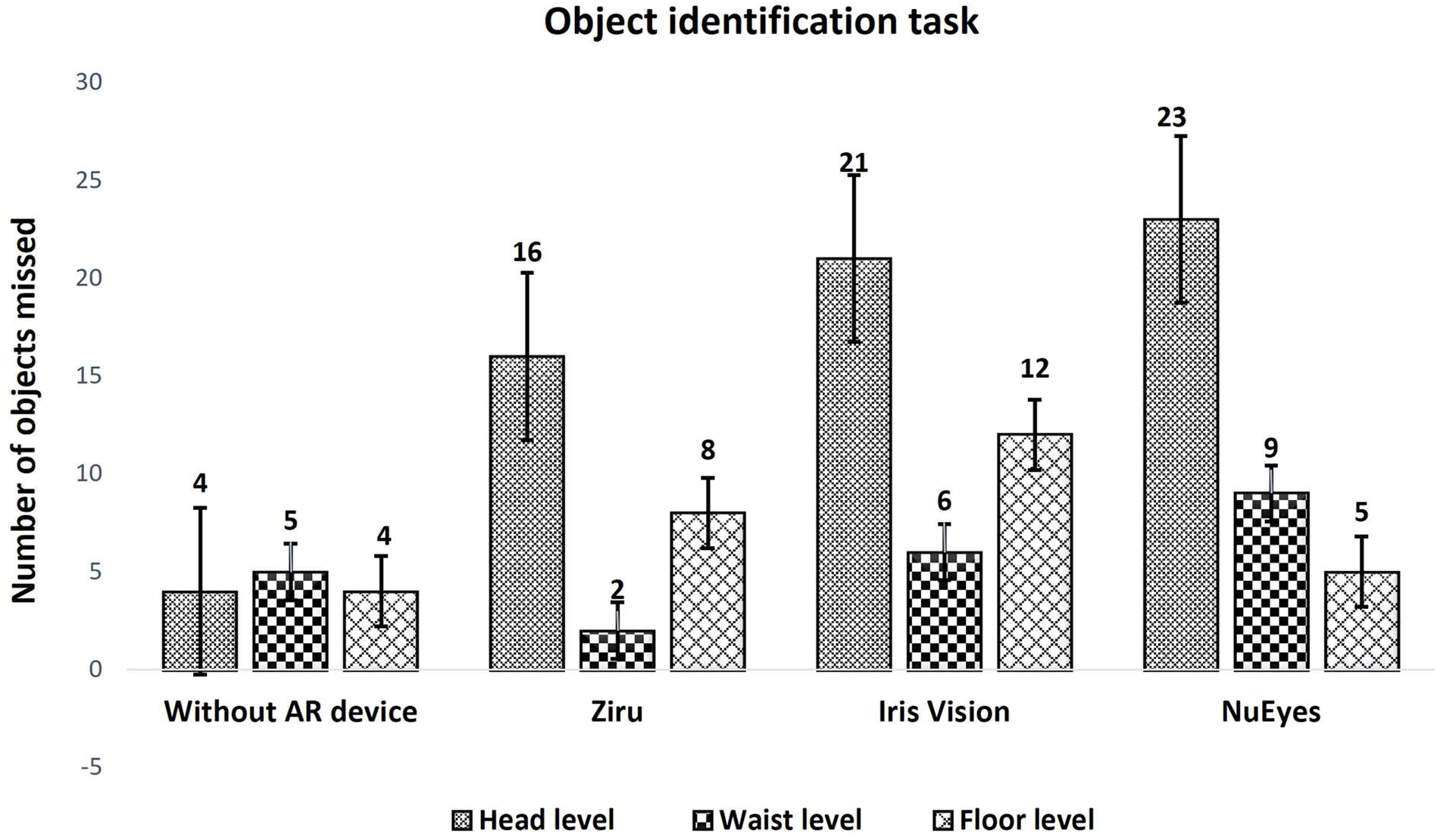
Object identification task performed without and with AR devices.

Fig 3: This figure demonstrates the number of objects missed at head level, waist level, and floor level. AR: Augmented reality.

## Discussion

Many studies have reported a change in the visual functioning of people with low vision using conventional optical low vision devices (LVD) and electronic devices^[7,8]^, however no study compared the performance of head-mounted displays (HMD). The augmented reality devices might replace the use of multiple low vision devices in the future. Augmented reality technology provides image enhancement that allows users to see the real world with enhanced brightness, magnification, contrast, and a combination of other elements^[8]^. Since many people with low vision have reduced contrast sensitivity, visual acuity, and/or loss of field of vision, it might be difficult or even impossible to have a “one size fits all” solution. With all the technological advances in the field of AR, it might provide more options for the visually impaired as explored in this study ^[21]^. There are a few studies that provide evidence that augmented reality devices could help improve visual acuity, contrast sensitivity, reading ability of people with low vision ^[22–25]^, however the studies aim at one or two visual functions which are not sufficient to conclude how these devices would be beneficial for performing daily living activities (functional vision). There is not enough literature comparing the visual functions of individuals with low vision while using different models of augmented reality (AR) devices, therefore this study highlights the comparison of three AR devices to determine what and how people with low vision could see with these devices. In addition, we have also assessed performance on a mobility task using all three devices.

All 27 participants were able to perform the visual task while using three models of AR devices, except that none of the participants were able to focus on small reading prints while using NuEyes, this implies that NuEyes cannot be used for reading fine prints. The majority of them were elderly people above 60 years of age, and most of them were found to have central field loss secondary to ocular pathology. Many studies have attempted to report the change in visual acuity of people with low vision while using AR devices^[16-18,21-23]^, yet this study compares the visual acuity in four different lighting conditions such as HLHC, HLLC, LLHC and LLLC. The change in the number of lines read was significantly different with Ziru and Iris Vision, but not with NuEyes in HLHC and HLLC condition. Interestingly, the change was significantly different with all three devices on low luminance high contrast (LLHC) and low luminance low contrast (LLLC) conditions which could be due to the nullifying effect of the background room illumination of 0.01lux. The display on the AR devices starts flickering only while viewing back illuminated charts at a certain level of magnification, which was not found while seeing externally illuminated charts or any other objects. This flickering could impede their performance at high magnification settings. For instance, the display started flickering from 3.5x level of magnification while using Ziru and it started from 2.5x level of magnification with IrisVision. Even though each magnification level of all the AR devices is relatively similar, the luminance level on the screen display plays a major role in visual perception. The majority of the participants were able to read the 5% low contrast chart using Ziru both on high luminance and on low luminance backgrounds which could be due to the higher luminance level of the screen display of Ziru. Interestingly, none of the participants were able to attempt reading the 5% contrast level chart with NuEyes. Most of them responded to 10% contrast level chart with IrisVision which implies that it was helpful seeing the medium level of contrast. Similar to many other studies^[18,21]^, we found a significant enhancement in near vision with Ziru and Iris Vision, but with NuEyes, the near vision reduced significantly which could be due to the focusing limitations of the device camera. Even though the NuEyes Pro 3 model was presented with an additional clip-on lens to focus farthest objects, it was not designed to read fine prints like newspapers, magazines, etc. Although, the mean distance and near contrast sensitivity improved significantly with Ziru and Iris Vision but it reduced significantly with NuEyes, this demonstrated that AR devices help improve the contrast sensitivity which is similar to previous studies ^[16,17,22,23]^. The luminance level was not cut down significantly while using Ziru compared to IrisVision and NuEyes, which could be the reason for participants reading more letters on distance and near contrast sensitivity chart with Ziru. The image quality of all the three devices can be visualized on the Supplementary Figure., it was obvious that the image quality was better in Ziru and IrisVision when compared to the NuEyes. We believe that the image quality was influenced by the camera resolution, image enhancement process, application properties, image processing methods and others for which further research is needed. Even though there were different contrast combination views available, all the participants preferred regular viewing mode while performing the visual tasks.

The depth perception test revealed that none of the AR devices helped in perceiving the depth between the boxes when compared to seeing without any AR device. Even though the mean correct responses showed a better score with Ziru when compared to the other two devices, none of the devices had a statistically significant difference. Similarly, except for three participants, no one was able to respond to the Frisby stereo plates. Previous reports explain that color vision has not been studied widely while using AR devices ^[21]^, in this study these AR devices were not noted to have any major difference in color and depth perception.

This is the first study to calculate and compare the functional vision score while using three AR devices. The functional acuity score (FAS) improved significantly with all three AR devices. However, the composite functional vision score (FVS) showed significant improvement only with Ziru and Iris Vision, it reduced significantly with NuEyes, which could be due to a notable reduction in the functional field score (FFS) with NuEyes. Even though the spectacle model AR device looks cosmetically appealing, and everyone prefers to choose the design when compared to the head mounted model, the field of vision and luminance has a significant reduction which is an inbuilt limitation of the spectacle model AR devices. Therefore, while we have enough evidence that magnification of font size is important for improving visual performance, it is also important to consider the field of view, contrast, brightness, and image quality of the devices before suggesting a device to people with low vision. Ophthalmologists, Optometrists, LVTs, OTs and people with low vision need to be cautious before choosing spectacle model AR devices for improving visual functions, especially for those with ocular conditions like retinitis pigmentosa, glaucoma and other ocular conditions that cause reduced field of vision.

The Htike et al study stated that there is no good evidence that HMDs improve walking efficiency^[19]^, while this study attempts to report a small component of walking during object identification task. During this task, all the participants were able to wear all three AR devices and move through the mobility course designed to identify objects at head level, waist level, and floor level including the signs for walking directions and distractors in the room. The number of objects missed was not significantly different while using Ziru among all three devices, which proved that there was no dampening of performance while using Ziru compared to without AR devices. The average time taken for identifying the tasks was lesser with IrisVision compared to condition seeing without AR devices and with other AR devices, which could be because of the participants were able to see the real world only through the device and there was no distraction outside their view. The number of objects missed was more with IrisVision and NuEyes when compared to Ziru.

Field of view of the AR devices and the type of visual field defects have important implications for visual performance using AR devices. Among the three devices tested, Ziru claims the largest field of view followed by IrisVision and NuEyes. It is interesting that the head level objects were missed more when compared to other levels with all three devices. The head level objects that were straight ahead, could have been hard to focus and identify by the majority of our participants who were older and with central field loss. Even though the task was performed at minimum level of magnification, still it would be a magnified scene when compared to seeing with naked eyes in real life scenario. So, it would have been easier for the participants to focus on waist-level and floor-level objects when compared to head-level objects. The participants had to move their head to fixate on any object of interest, while they tried to look at floor-level or waist-level objects, there was an adequate amount of distance for them to focus on the objects. Previous studies have reported that people with low vision spend a higher cognitive load while walking, which may affect their ability to visually process the displayed content^4^, therefore further research is needed to explore how people identify objects by compensating the magnification level of AR devices in a real-world environment. However, this study reveals that it might be possible to utilize AR devices for identifying objects while navigating in an indoor environment.

Apart from the potential of AR devices in general which has been established already by many researchers, using these devices as accessibility tools has many advantages including distance viewing, reading, contrast enhancement, etc. The cost of AR devices differs a lot widely, however, it is important to consider the performance of the device for improving the visual functions of people with low vision. Surprisingly, Ziru which costs $700 functions equally well and sometimes better when compared to IrisVision $2999 or NuEyes $3999, which brings in a hope of availability of AR devices at very affordable cost.

In this study, we have used a heterogeneous group of individuals with a wide range of ocular conditions. While such a group is representative of the general low vision population, the wide variety of ocular pathologies and the limited sample size prevent drawing further conclusions about which type of AR devices are more effective for specific ocular conditions. However, both stand-alone and head-mounted AR models were found to be more effective in improving visual function compared to spectacle model AR devices. The time taken for exploring the AR devices is a limitation in this study which is similar to findings from previous studies, people with low vision need adequate time to explore these devices which would be helpful in receiving more feedback. However, further follow-up study is recommended where the AR devices can be given to the participants for a home loan to understand their perception about the devices. Further research can explore how these findings can be applied to improve the home and work environment of people with low vision. Of note, in this study we compared object detection using the three AR devices while the participants were performing a mobility task. Even though HMDs are not recommended and presents safety concerns for use while moving since the magnification and limited field of view can interfere with distance judgements and perception, we found that our participants benefitted from using the devices for detecting and identifying objects during mobility. The settings used for the mobility task in this study are different from less controlled real-world settings and therefore, further research is needed before extending the use to real-world. However, HMDs are becoming increasingly popular as AR/VR based assistive technologies are being developed for various activities of daily life that includes navigation and wayfinding^[4,27-29].^

## Conclusion

The majority of the visual functions had improvement with all three devices, yet the distance vision had the most enhancement in all the illumination settings using the Ziru and IrisVision, whereas the enhancement was limited to certain lighting conditions with NuEyes. The near visual acuity, distance contrast sensitivity, and near contrast sensitivity had improvement with Ziru and IrisVision, but not with NuEyes. The functional vision score improved with Ziru and IrisVision significantly, whereas it reduced significantly with NuEyes. While there are few changes in visual functions with all three models of AR devices, there was a huge difference in terms of cost. Researchers and individuals with low vision expect that augmented reality (AR) devices in the form of spectacles would be a good option; however, this expectation was not supported by the study in terms of improving visual functions. The study found that stand-alone and head-mounted AR models were more effective in improving visual functions than spectacle models. While augmented reality devices do help in enhancing visual performance, it is important for them to have a simple design with the most essential functions.

## Data Availability

The datasets generated and/or analyzed during the current study are available from the corresponding author upon reasonable request.

---

**Supplementary figure.**
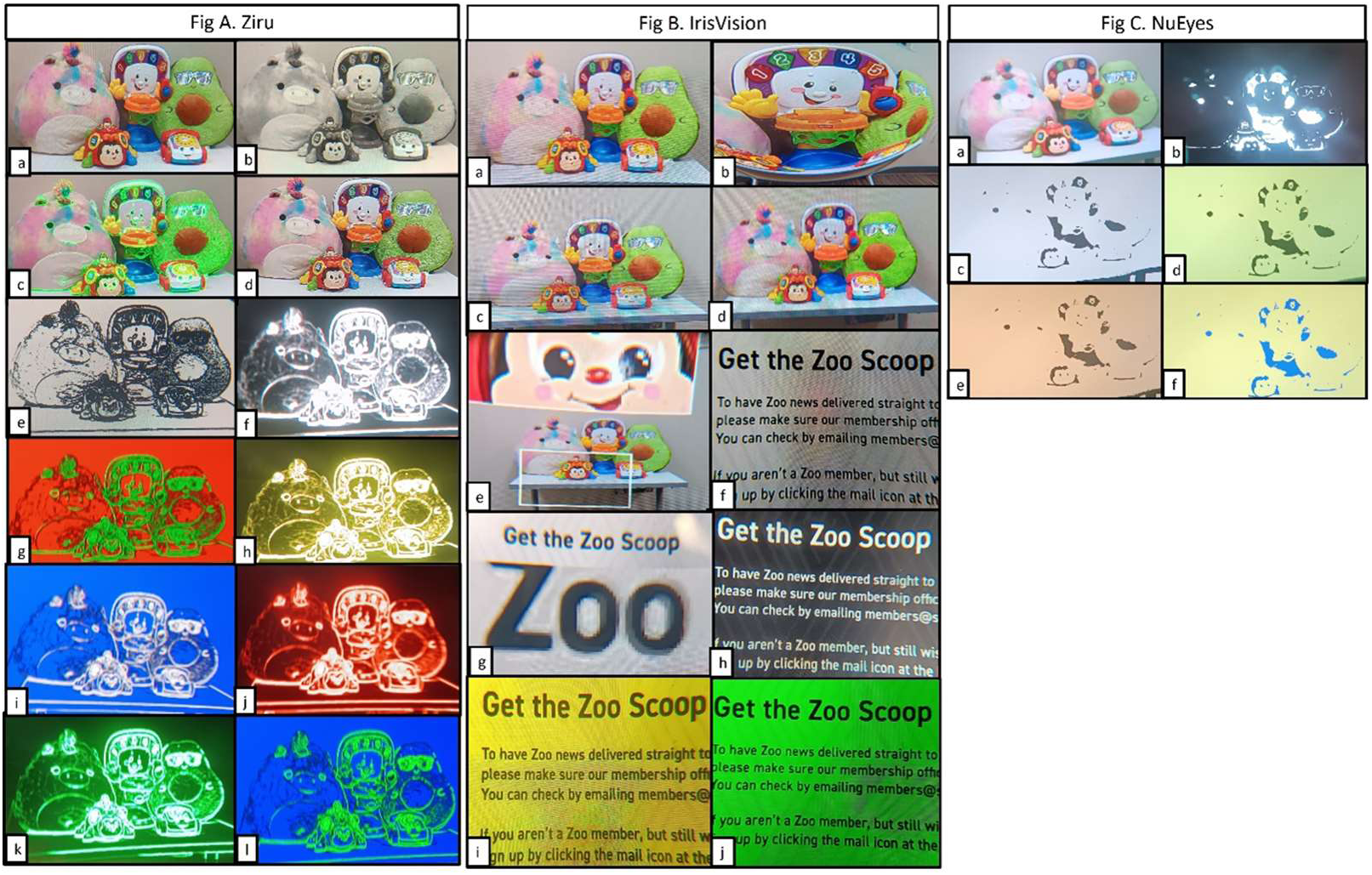
Graphical representation of different viewing modes of Ziru, IrisVision and NuEyes. Figure A. Ziru. a. Clear, b. Gray, c. Boost, d. Cogni, e. Outline - Black Outline white background, f. White outline black background, g. Green outline red background, h. Yellow outline black background, i. White outline blue background, j. Red outline black background, k. Green outline black background, l. Green outline blue background. Figure B. IrisVision. a. Scene, b. Scene with bubble, c. RP, d. Television, e. Bioptic, f. Reading, g. Reading line, h. Reading inverted, i. Reading yellow, j. Reading green. Figure C. NuEyes. a. Regular, b. Black, c. White, d. Yellow, e. Orange, f. Blue

## References

1. Thylefors B, Négrel AD, Pararajasegaram R, Dadzie KY. Global data on blindness. Bulletin of the world health organization. 1995;73(1):115.

2. Flaxman AD, Wittenborn JS, Robalik T, Gulia R, Gerzoff RB, Lundeen EA, Saaddine J, Rein DB, Baldonado KN, Davidson C, Dougherty MC. Prevalence of visual acuity loss or blindness in the US: a Bayesian meta-analysis. JAMA Ophthalmol. 2021 Jul 1;139(7):717–23.

3. Rein DB, Wittenborn JS, Zhang P, Sublett F, Lamuda PA, Lundeen EA, Saaddine J. The economic burden of vision loss and blindness in the United States. Ophthalmology. 2022 Apr 1;129(4):369–78.

4. Zhao Y, Hu M, Hashash S, Azenkot S. Understanding low vision people’s visual perception on commercial augmented reality glasses. Conf. Hum. Factors Comput. Syst. 2017 May 2 (pp. 4170–4181).

5. Abdulhussein D, Abdul Hussein M. WHO Vision 2020: Have we done it?. Ophthalmic Epidemiol. 2023 Jul 4;30(4):331–9.

6. Kartha A, Sadeghi R, Bradley C, Tran C, Gee W, Dagnelie G. Measuring visual information gathering in individuals with ultra low vision using virtual reality. Sci. Rep. 2023 Feb 23;13(1):3143.

7. Gopalakrishnan S, Paramasivan G, Sathyaprasath M, Raman R. Preference of low vision devices in patients with central field loss and peripheral field loss. Saudi J Ophthalmol. 2021 Oct 1;35(4):286–92.

8. Deemer AD, Bradley CK, Ross NC, Natale DM, Itthipanichpong R, Werblin FS, Massof RW. Low vision enhancement with head-mounted video display systems: are we there yet?. Optom. Vis. Sci. 2018 Sep 1;95(9):694–703.

9. Massof RW, Fletcher DC. Evaluation of the NEI visual functioning questionnaire as an interval measure of visual ability in low vision. Vis. Res. 2001 Feb 1;41(3):397–413.

10. Garcia GA, Khoshnevis M, Gale J, Frousiakis SE, Hwang TJ, Poincenot L, Karanjia R, Baron D, Sadun AA. Profound vision loss impairs psychological well-being in young and middle-aged individuals. Clin Ophthalmol. 2017 Feb 22:417–27.

11. Wolffsohn JS, Cochrane AL. Design of the low vision quality-of-life questionnaire (LVQOL) and measuring the outcome of low-vision rehabilitation. Am. J. Ophthalmol. 2000 Dec 1;130(6):793–802.

12. Evans JR, Fletcher AE, Wormald RP. Depression and anxiety in visually impaired older people. Ophthalmology. 2007 Feb 1;114(2):283–8.

13. Deemer AD, Swenor BK, Fujiwara K, Deremeik JT, Ross NC, Natale DM, Bradley CK, Werblin FS, Massof RW. Preliminary evaluation of two digital image processing strategies for head-mounted magnification for low vision patients. Transl. Vis. Sci. Technol. 2019 Jan 2;8(1):23.

14. Peli E, Luo G, Bowers A, Rensing N. Development and evaluation of vision multiplexing devices for vision impairments. Int. J. Artif. Intell. 2009 Jun;18(03):365–78.

15. Cheng D, Wang Q, Liu Y, Chen H, Ni D, Wang X, Yao C, Hou Q, Hou W, Luo G, Wang Y. Design and manufacture AR head-mounted displays: A review and outlook. Light: Adv. Manuf. 2021 Sep 18;2(3):350–69.

16. Kinateder M, Gualtieri J, Dunn MJ, Jarosz W, Yang XD, Cooper EA. Using an augmented reality device as a distance-based vision aid-promise and limitations. Optom.Vis. Sci. 2018 Sep 1;95(9):727–37.

17. Culham LE, Chabra A, Rubin GS. Clinical performance of electronic, head-mounted, low-vision devices. Ophthal. Physiol. Opt. 2004 Jul;24(4):281–90.

18. Miller A, Crossland MD, Macnaughton J, Latham K. Are wearable electronic vision enhancement systems (wEVES) beneficial for people with age-related macular degeneration? A scoping review. Ophthal.Physiol. Opt. 2023 Jul;43(4):680–701.

19. Min Htike H, H. Margrain T, Lai YK, Eslambolchilar P. Augmented reality glasses as an orientation and mobility aid for people with low vision: a feasibility study of experiences and requirements. Conf. Hum. Factors Comput. Syst. 2021 May 6 (pp. 1–15).

20. Fuhr P, Holmes L, Fletcher D, Swanson M, Kuyk T. The AMA Guides functional vision score is a better predictor of vision-targeted quality of life than traditional measures of visual acuity or visual field extent. Vis. Impair. Res. 2003 Jan 1;5(3):137–46.

21. Li Y, Kim K, Erickson A, Norouzi N, Jules J, Bruder G, Welch GF. A scoping review of assistance and therapy with head-mounted displays for people who are visually impaired. ACM Trans. Access. Comput. 2022 Aug 19;15(3):1–28.

22. Yeo JH, Bae SH, Lee SH, Kim KW, Moon NJ. Clinical performance of a smartphone-based low vision aid. Sci. Rep. 2022 Jun 24;12(1):10752.

23. Wittich W, Lorenzini MC, Markowitz SN, Tolentino M, Gartner SA, Goldstein JE, Dagnelie G. The effect of a head-mounted low vision device on visual function. Optom. Vis. Sci. 2018 Sep 1;95(9):774–84.

24. Gopalakrishnan S, Suwalal SC, Bhaskaran G, Raman R. Use of augmented reality technology for improving visual acuity of individuals with low vision. Indian J. Ophthalmol. 2020 Jun 1;68(6):1136–42.

25. Crossland MD, Starke SD, Imielski P, Wolffsohn JS, Webster AR. Benefit of an electronic head-mounted low vision aid. Ophthal.Physiol. Opt. 2019 Nov;39(6):422–31.

26. Fox DR, Ahmadzada A, Friedman CT, Azenkot S, Chu MA, Manduchi R, Cooper EA. Using augmented reality to cue obstacles for people with low vision. Opt. Express. 2023 Feb 9;31(4):6827–48.

27. Hamilton-Fletcher G, Liu M, Sheng D, Feng C, Hudson TE, Rizzo JR, Chan KC. Accuracy and Usability of Smartphone-Based Distance Estimation Approaches for Visual Assistive Technology Development. IEEE OJEMB. 2024 Jan 25.

28. Ricci FS, Boldini A, Ma X, Beheshti M, Geruschat DR, Seiple WH, Rizzo JR, Porfiri M. Virtual reality as a means to explore assistive technologies for the visually impaired PLOS Digit Health. 2023 Jun 20;2(6):e0000275.

29. Ricci FS, Boldini A, Beheshti M, Rizzo JR, Porfiri M. A virtual reality platform to simulate orientation and mobility training for the visually impaired. Virtual Real. 2023 Jun;27(2):797–814.

